# Maternal HIV Infection and Low Birthweight Outcomes among Children born to Adolescent and Young Adult Mothers in Zambia: A Multi-level Analysis

**DOI:** 10.1101/2025.10.21.25338507

**Authors:** Womba Samudimu, Samson Shumba

## Abstract

Infants born with LBW are at a higher risk of dying within their first month of life, and those who survive are prone to lifelong challenges, including stunted growth, lower IQ, and an increased risk of chronic conditions in adulthood, such as obesity and diabetes. Infants with LBW may have digestive and breathing problems and complications in eating, gaining weight, and fighting off infections compared with normal birth weight infants. The aim of this study was to find out the effects of maternal HIV infection on low birthweight among children born to adolescents and young adult mothers in Zambia.

The study used a cross-sectional study design using the Zambia demographic health survey which was conducted in all the 10 provinces. The study population comprised of children born to adolescent and young adult mothers five years preceding the survey. Overall, LBW prevalence did not differ significantly by HIV status, with HIV-positive mothers recording 9.6% and HIV-negative mothers 8.7%. Provincial variation was observed, with LBW among HIV-infected mothers highest in Eastern (15.0%) and Northern (14.2%) and lowest in Muchinga (0%) and Luapula (1.6%). Among non-infected mothers, Lusaka (10.7%) and Muchinga (10.0%) reported the highest LBW prevalence, while Northern had the lowest (3.9%). A multi-level analysis confirmed that maternal HIV status was not significantly associated with LBW (OR = 0.95, 95% CI: 0.50–1.79, p = 0.871). Instead, LBW was strongly predicted by socio-economic and maternal factors: mothers in the middle wealth index had higher odds of LBW (OR = 1.69, 95% CI: 1.16–2.47, p = 0.006), home deliveries had reduced odds (OR = 0.28, 95% CI: 0.14–0.59, p = 0.001), and multiple births increased the odds more than eight-fold (OR = 8.05, 95% CI: 3.64–17.83, p < 0.001).

The study indicates that HIV infection alone may not be a principal factor influencing LBW outcomes in this age group in Zambia rather, LBW is more strongly shaped by socio-economic conditions, delivery setting, and multiple birth risks. These results are essential for shaping future HIV/AIDS interventions, guidelines, and policies designed to avert LBW in at-risk populations.

## Background

Low birth weight (LBW) is a major public health concern and a critical risk factor for perinatal survival, as well as infant and child mortality and morbidity in early childhood (ages 3–8 years). It significantly contributes to the global burden of infant mortality [1]. The World Health Assembly Resolution 65.6, set a targets to reduce LBW by 30% by 2025 [2]. This would result in a 3% annual relative decrease from 2012 to 2025, leading to a drop of around 20 million to nearly 14 million newborns with low birth weight[3].Sub-Saharan Africa (15%) and South-central Asia (27%) regions share a disproportionate burden of LBW [4]. In Zambia, about 11% of newborns in Lusaka were LBW [4]. HIV-positive women are twice as likely to deliver LBW infants as HIV-negative women. Zambia, a LMIC, is likely to have women unable to meet the nutritional demands of pregnancy due to HIV infection [5]. According to the Zambia demographic health survey, 9% of births were reported to be smaller than average, and 2% as very small[6].

Infants born with LBW are at a higher risk of dying within their first month of life, and those who survive are prone to lifelong challenges, including stunted growth, lower IQ, and an increased risk of chronic conditions in adulthood, such as obesity and diabetes (UNICEF, 2023). Infants with LBW may have digestive and breathing problems and complications in eating, gaining weight, and fighting off infections compared with normal birth weight infants. It has been estimated that over 20 million deliveries have resulted in LBW infants (LBW; <2,500 g) annually; the vast majority of these LBW deliveries are in low- and middle-income countries [7]. Several factors, such as infections, low education level, low income, and occupation, are associated with LBW [7]. Mothers who had given a female neonate had three times risk of LBW than those who gave a male baby [8]. In addition, one or more antenatal care (ANC) visits were associated with reduced odds of LBW compared to those who had no follow-ups [8].

Infants born to HIV-infected women have been reported also to be thinner, with smaller head circumferences than infants of HIV-negative mothers [9]. Quality of pregnancy outcomes associated with use Anti-Retroviral Therapy (ART), in HIV patients receiving ART have reported lower cases of low birth weight and premature weight [10]. Therefore, attempts to determine a comparative analysis of low birthweight among HIV infected and non-infected adolescent and young adult mothers in Zambia is needed hence the aim of this study. This study is imperative because it will determine the direction of policy making regarding HIV and low birth weight in Zambia.

Low birth weight (LBW) is influenced by maternal, socio-economic, and healthcare factors. In Zambia, approximately 1.2% of female adolescents aged 10–14 and 1.8% of those aged 15–19 are HIV-positive, and maternal HIV infection has been linked to higher LBW risk[11] Other key determinants include preeclampsia, preterm birth, multiple pregnancies, adolescent age, low maternal education, poor household wealth, and inadequate antenatal care [12].

Given this dual burden of HIV and LBW, it is essential to examine the relationship between maternal HIV status and birth outcomes among adolescents and young adults. Such analysis not only enhances understanding of the determinants of LBW but also informs the design and strengthening of HIV/AIDS interventions, guidelines, and policies targeting vulnerable populations. The aim of this study was to investigate the association between maternal HIV infection and LBW among adolescent and young adult mothers in Zambia, while also assessing the role of socio-demographic, maternal, and contextual factors using multilevel analysis.

### Conceptual Framework

Figure 1 shows the conceptual framework was adopted from WHO and UNICEF showing various factors that are associated with low birthweight [13].

**Figure 1:**
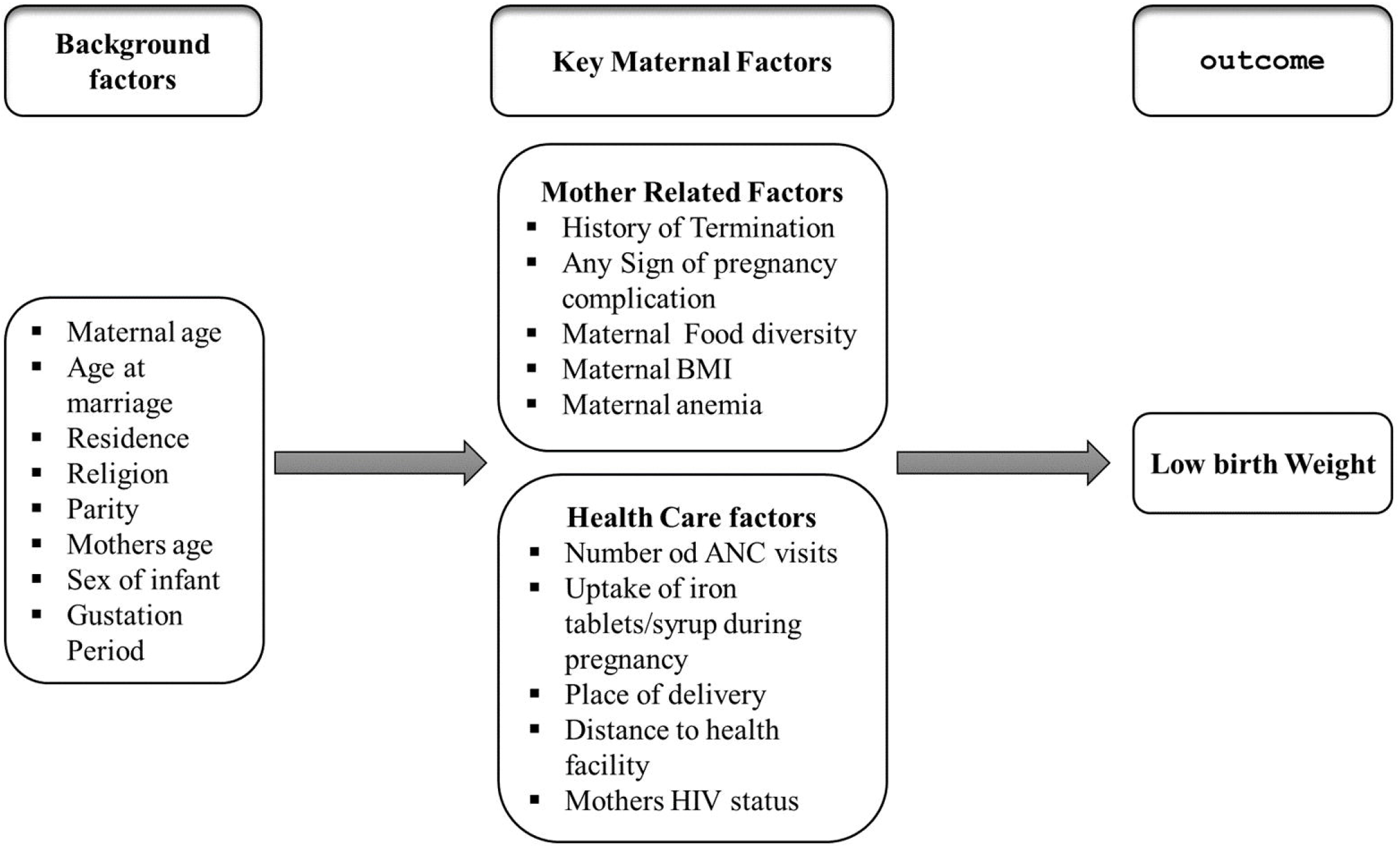
Conceptual Framework of Factors Associated with Low Birthweight. Conceptual framework.

## Methods

### Study Design, study population and inclusion criteria

The study used a cross-sectional study design using the Zambia demographic health survey is cross sectional study which was conducted in all the 10 provinces. The survey collected data on levels of fertility, maternal mortality, maternal and child health, contraceptive use, infant, child and neonatal mortality and HIV prevalence among the reproductive age group for men and women. The 2018 ZDHS followed a stratified two-stage sample design. The first stage involved selecting sample points (clusters) consisting of EAs. EAs were selected with a probability proportional to their size within each sampling stratum.

The study population comprised of adolescent mothers who gave birth to live babies five years preceding the survey. All adolescent mothers who gave birth to live babies without birth weight will be excluded from the study. The study will also include women who know their HIV status and those who do not know will be excluded as shown in Fig 2.

**Figure 2:**
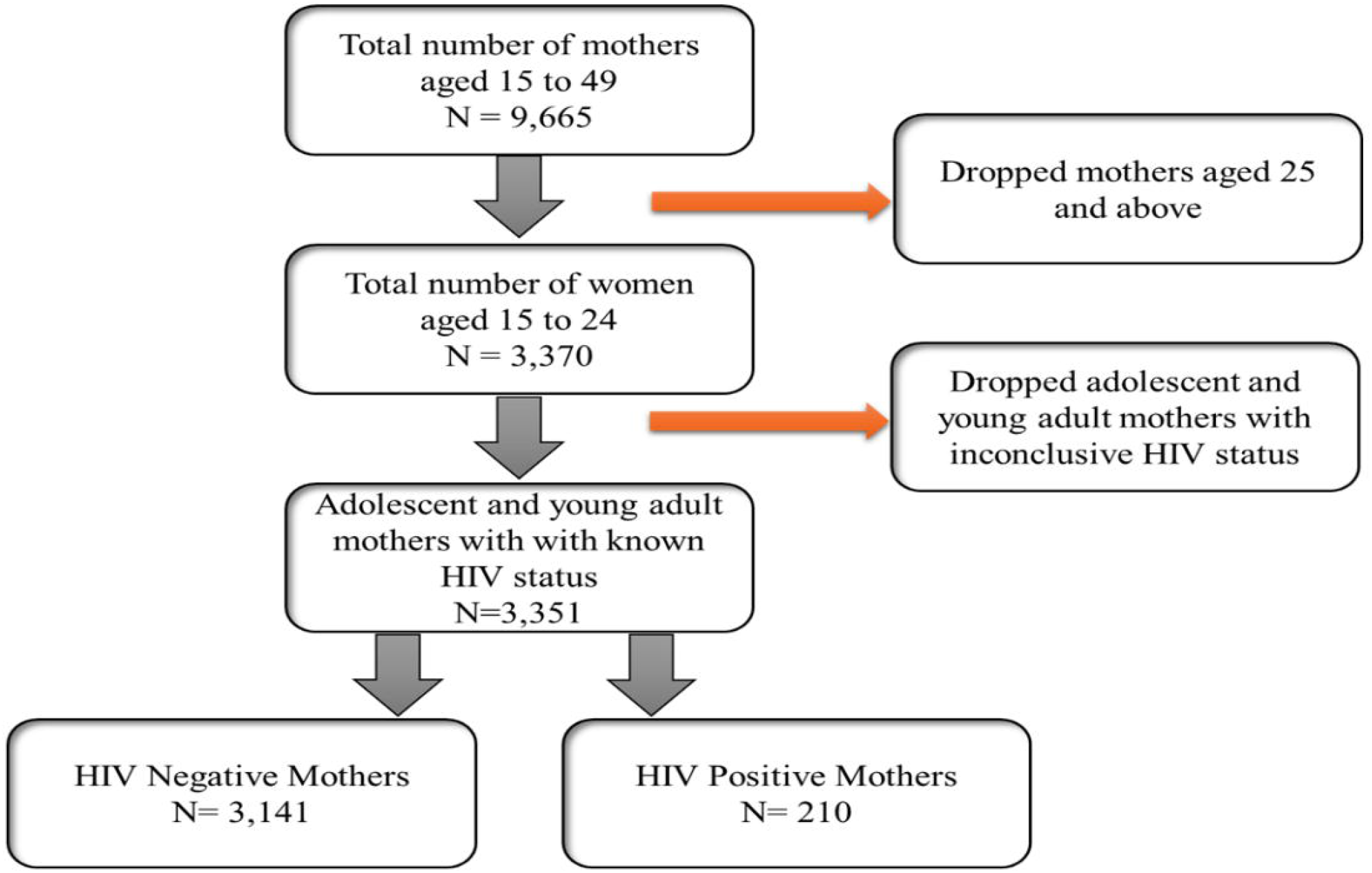
Description Sample derivation criteria. Overview of Sample Derivation Criteria.

**Figure 3:**
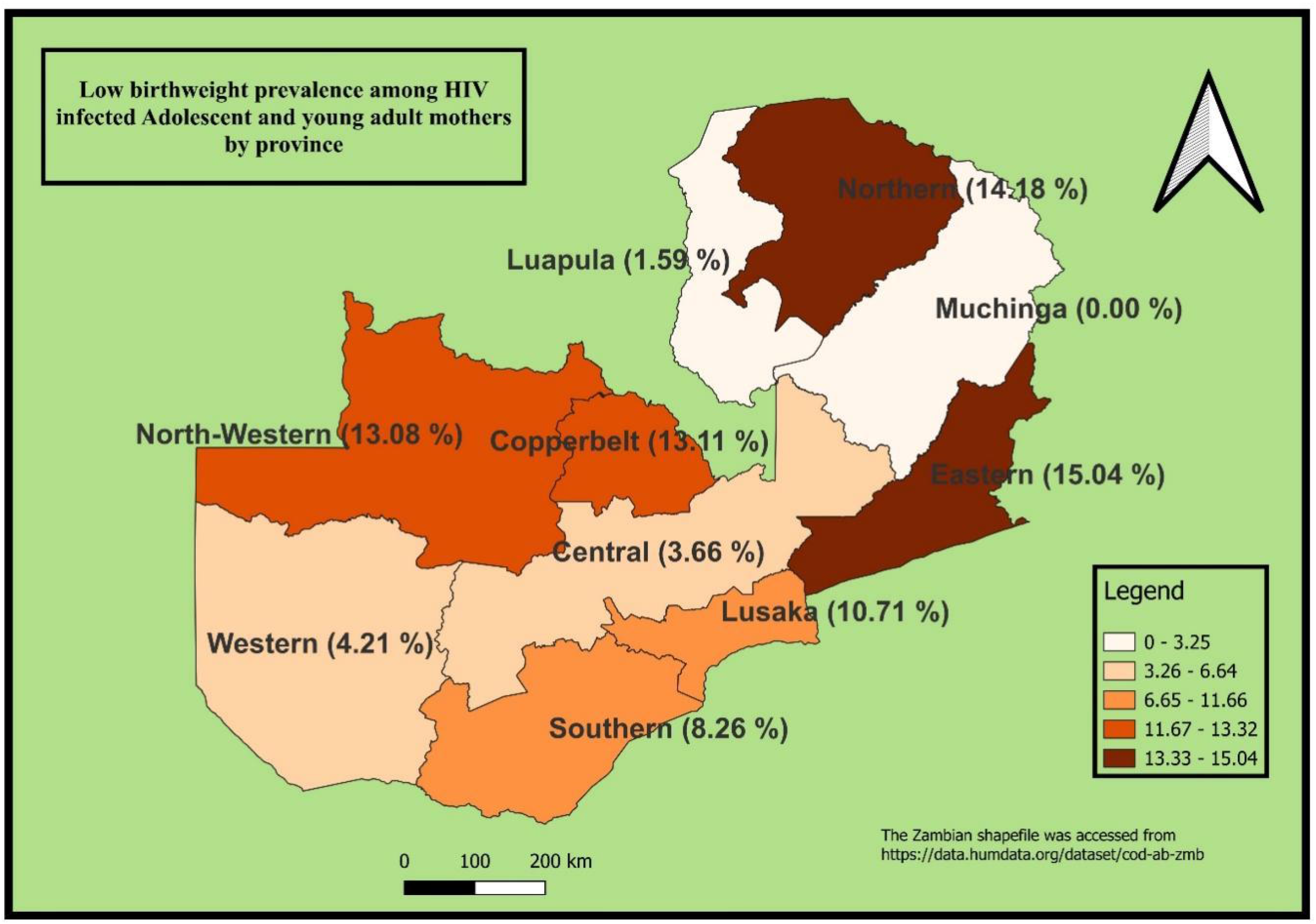
Geographic Distribution of Low Birthweight Prevalence among HIV-Infected Adolescent and Young Adult Mothers by Province, Zambia. The maps were generated using QGIS version 3.4.1. (Source: author generated).

**Figure 4:**
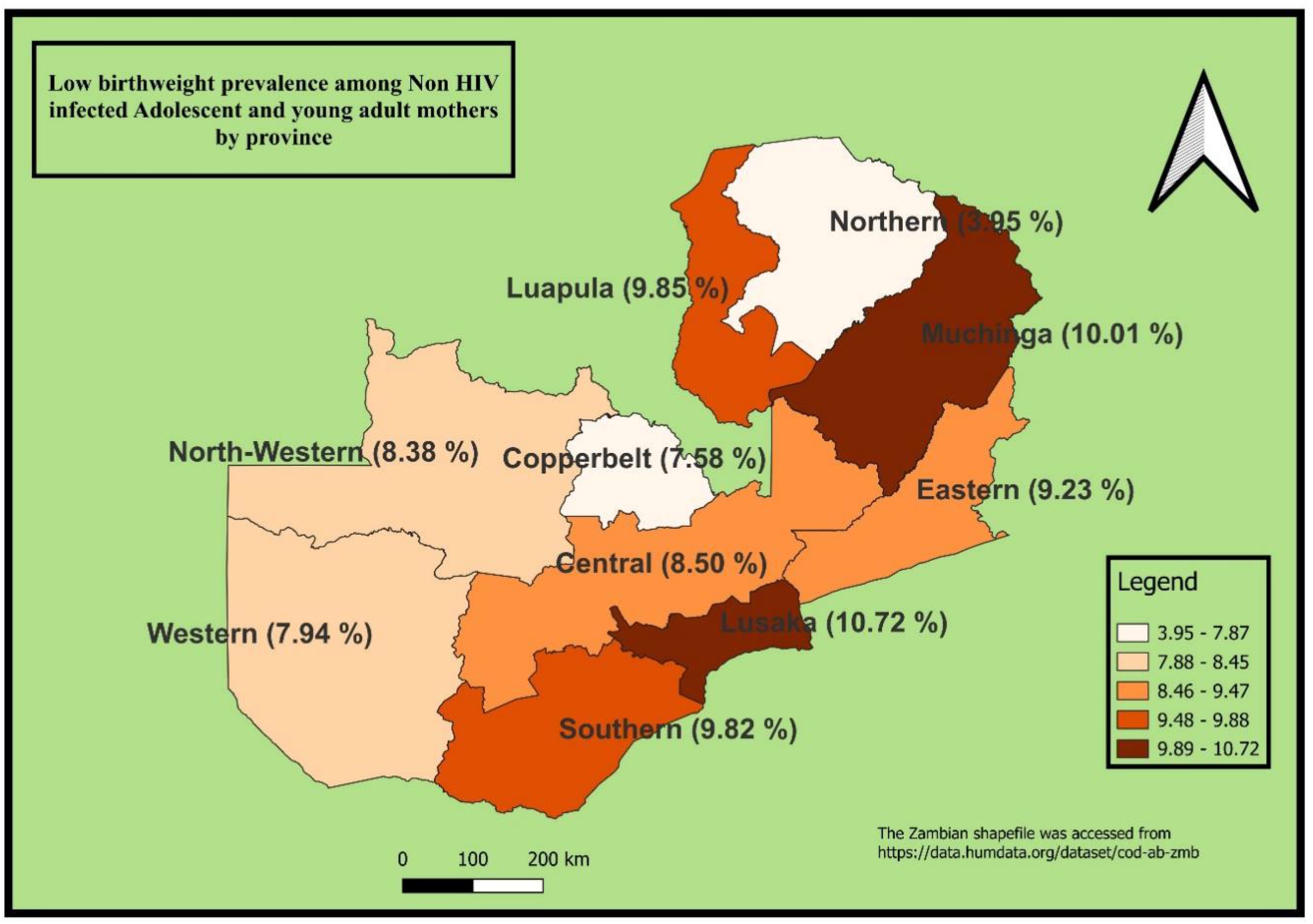
Geographic Distribution of Low Birthweight Prevalence among HIV-Infected Adolescent and Young Adult Mothers by Province, Zambia. The maps were generated using QGIS version 3.4.1. (Source: author generated).

For this study, relevant variables were extracted from the children’s data files of the 2018 ZDHS dataset. The analysis focused specifically on adolescent and young adult mothers aged 15 to 24 years, with women aged 25 years and older excluded from the study sample.

### Individuals (Dependent and Independent Variables) and community level factors

The variable of interest in this study is low birth weight. Background variables included in the above conceptual are maternal age, age at marriage, residence (rural or urban), parity, mothers age, sex of infant, gestation period. Age was treated as a continuous variable. Sex of infant was categorized as either male or female.

Community-level variables in this study were created by clustering individual-level data and included community poverty, education, knowledge of family planning (FP) methods, and place of residence. These variables were categorized as ‘low’ or ‘high,’ representing the prevalence of each characteristic within the clusters. The variables for place of residence and geographic region were retained in their original forms, with place of residence being critical in the sample design, used to estimate key demographic and health indicators at the national level. It was classified as ‘rural’ or ‘urban,’ contributing significantly to the characterization of the communities.

### Data Analysis

The study provides descriptive statistics, including frequencies and percentages for categorical variables, adjusted for sample weighting to ensure accurate representation. To examine the association between the outcome variable (low birthweight, coded as 0 for “2500g and above” and 1 for “less than 2500g”) and the categorical predictors, the uncorrelated Design-Based Chi-square test (Rao-Scott Chi-square test) was applied. The study further employed Mixed Effect Logistic Regression to account for the hierarchical structure of the data, with adolescent and young adult mothers nested within households, and households nested within clusters. Variables included in the model were selected based on comprehensive literature review, adhering to an investigator-led approach. Model selection was guided by evaluating the Akaike Information Criterion (AIC), and Bayesian Information Criterion (BIC) to identify the best fit. Multicolinearity was assessed, and the variance of predictors was found to be less than 5, indicating no significant concern. Data analysis was conducted using Stata version 14.2.

### Model Specification of the Mixed Effect Logistic Regression

Let *Y*_*ij*_represent the binary outcome variable for the *i*− *th* individual within the *j*-*th* cluster (districts). The outcome *Y*_*ij*_ is coded as 1 if low birthweight (less than 2500g) and 0 otherwise. The mixed-effect logistic regression model can be specified as (Fitzmaurice et al, 2012; Rabe-Hesketh and Skrondal, 2012).

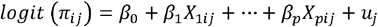

Where:

- 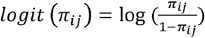 is the log odds of the outcome for the *i*-*th* Individual in the *j*-*th* cluster.
- *π*_*ij*_ Is the probability of the outcome when all predictors are zero.
- *α*_1_, *α*_2_,… *α*_*p*_, are the fixed coefficients for the covariates *X*_1*ij*_, *X*_2*ij*_,…, *X*_*pij*_, which are individual and community level predictors such as age, education, occupation etc.
- *u*_*j*_ represents the random effect associated with cluster *j* (cluster number), assumed to be normally distributed with mean zero and variance 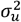. This random effect captures the unobserved heterogeneity at the cluster level.

This model allows for the inclusion of both fixed effects, accounting for individual and community-level factors, and random effects, addressing the hierarchical structure of the data. The use of mixed-effect logistic regression provides robust estimates by accounting for clustering of adolescent and young adult mothers within households and households within clusters.

### Ethical Consideration

The 2018 Zambia Demographic and Health Survey (ZDHS) received ethical clearance from the ICF Institutional Review Board (IRB) under Project Number 132989.0.000.ZM.DHS.02. Permission to access and utilize the ZDHS dataset (ZMKR71FL.DTA) was obtained from ICF Macro, with the data available at The DHS Program. The analysis strictly adhered to the survey’s data use guidelines, prioritizing data confidentiality and ensuring that no attempt was made to identify any households or individual respondents, thus maintaining participant anonymity.

## Results

### Bivariate Analysis of Factors Associated with Low Birth Weight

The study investigated the association between maternal and household characteristics and the likelihood of low birth weight (LBW) among adolescent and young adult mothers in Zambia. Overall, four factors were significantly associated with LBW: wealth index, media exposure, place of delivery, and multiple births. Mothers from middle-income households had the highest prevalence of LBW (13.2%), a surprising trend compared to poor (7.24%) and rich households (8.76%). Additionally, multiple births showed a markedly higher LBW rate (44.84%) compared to singletons (8.06%).

Place of delivery was also a strong predictor of LBW. Births in private health facilities had the highest proportion of LBW (13.95%), followed by public facilities (9.26%), while home deliveries had the lowest rate (2.49%). Other variables, such as maternal age, HIV status, residence, employment, and antenatal care attendance, were not significantly associated with LBW in this bivariate analysis. However, marital status approached statistical significance (p=0.054), suggesting a potential protective effect of being married.

**Table 1:**
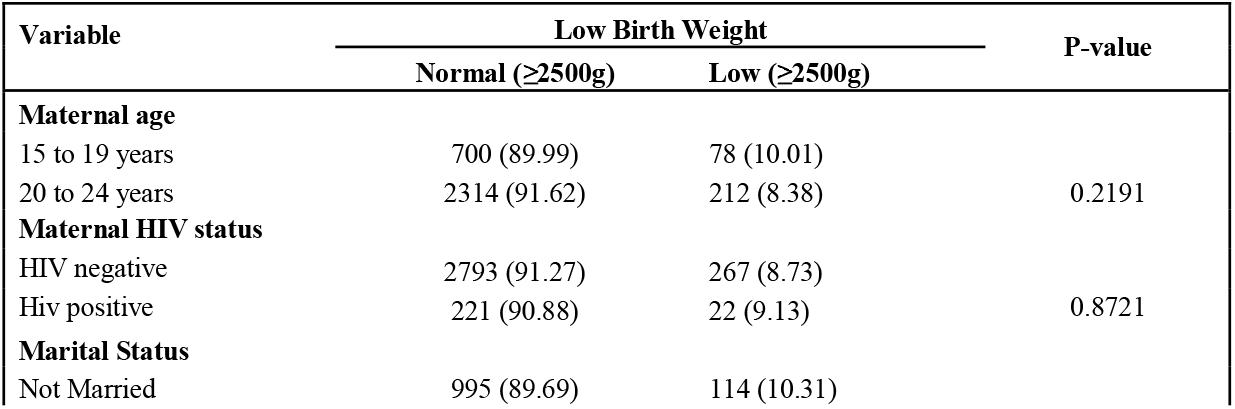
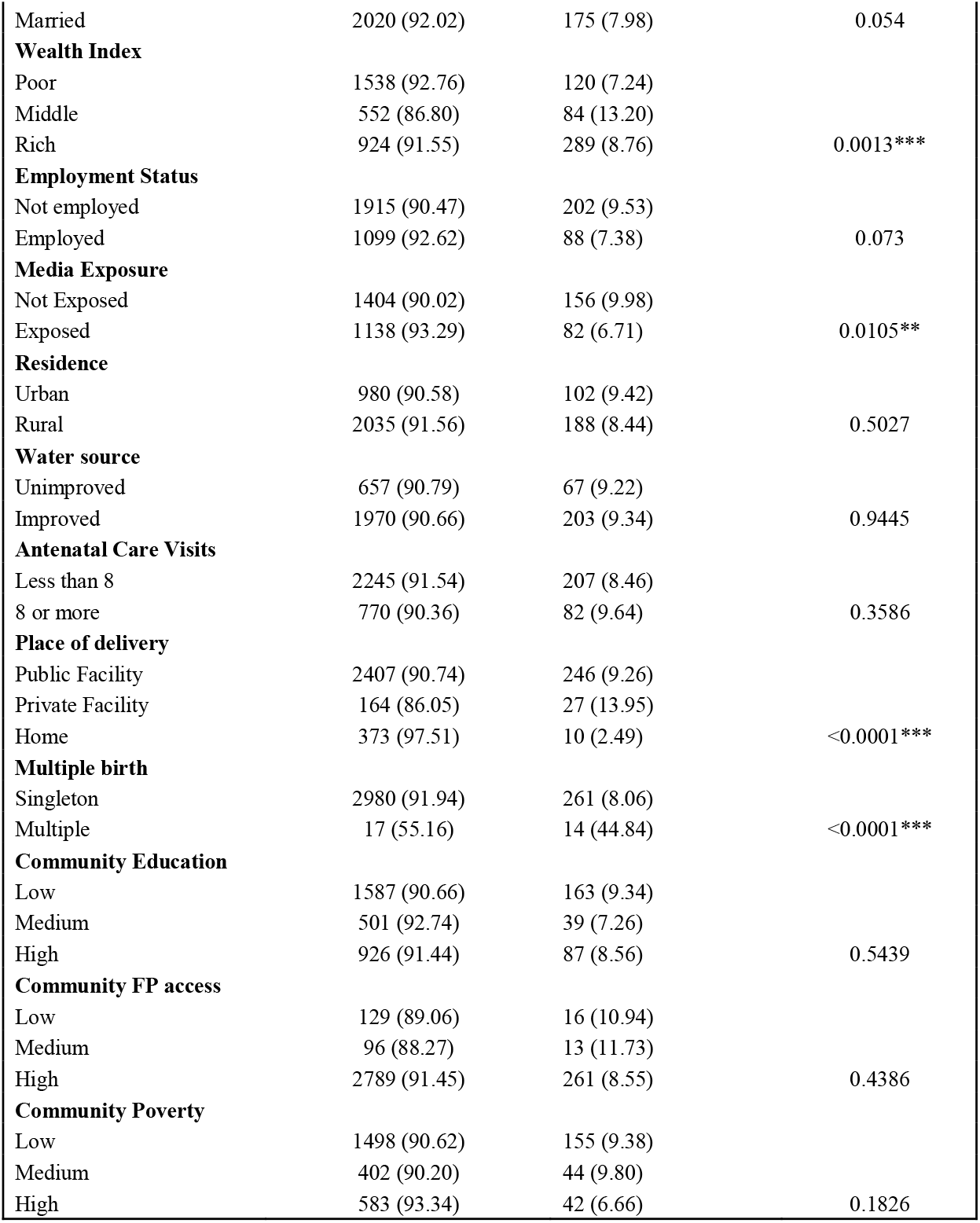
Distribution of Low Birth Weight by Maternal and Household Characteristics among Adolescent and Young Adult Mothers in Zambia.

### Provincial Prevalence of Low Birthweight among HIV-Infected Adolescent and Young Adult Mothers in Zambia

The prevalence of low birthweight among HIV-infected adolescent and young adult mothers shows substantial spatial variation across Zambia’s provinces. Eastern Province (15.04%) and Northern Province (14.18%) recorded the highest LBW rates. Similarly, Copperbelt (13.11%) and North Western (13.08%) also had elevated LBW prevalence, suggesting a need for targeted maternal health interventions in these areas. In contrast, some provinces showed remarkably low LBW prevalence, such as Muchinga (0%), Luapula (1.59%), and Central (3.66%), possibly indicating better outcomes or data limitations (e.g., small sample sizes). Lusaka, the capital, reported a moderate prevalence of 10.71%, while Southern and Western provinces had intermediate levels at 8.26% and 4.21%, respectively.

### Provincial Prevalence of Low Birthweight among Non-HIV-Infected Adolescent and Young Adult Mothers in Zambia

The data presents the prevalence of low birthweight among non–HIV-infected adolescent and young adult mothers across Zambia’s provinces. The figures vary notably by region, with Lusaka recording the highest prevalence at 10.72 percent, followed by Muchinga (10.01 percent), Luapula (9.85 percent), and Southern Province (9.82 percent). These elevated rates may reflect challenges such as limited access to quality antenatal care, poor maternal nutrition, and a higher incidence of adolescent pregnancies. Eastern Province also shows a relatively high prevalence at 9.23 percent, while Central and North Western provinces are closer to average, reporting 8.5 percent and 8.38 percent, respectively. Copperbelt and Western provinces show slightly lower prevalence rates at 7.58 percent and 7.94 percent. Notably, Northern Province records the lowest prevalence at 3.95 percent.

### Multilevel mixed effect logistic regression of low birthweight neonates of HIV-infected and non-infected adolescent and young adult mothers

The mixed-effects logistic regression model assessed the association between maternal and contextual factors and the likelihood of low birthweight among adolescent and young adult mothers in Zambia, accounting for clustering at the enumeration area level. The analysis revealed that most maternal socio-demographic variables such as age, education, marital status, employment, media exposure, and HIV status were not significantly associated with low birthweight. For example, maternal HIV-positive status did not significantly predict low birthweight (OR = 0.95, 95% CI: 0.50–1.79, p = 0.871), suggesting that HIV status alone may not substantially influence birthweight outcomes in this population. Similarly, being in the 20–24 age group was associated with lower odds of low birthweight compared to adolescents aged 15–19 (OR = 0.78, 95% CI: 0.56–1.09), although this difference was not statistically significant (p = 0.146), indicating no clear age-related variation within this narrow age band.

However, several variables showed significant associations. Women in the middle wealth index had significantly higher odds of delivering a low birthweight baby compared to those in the poorest group (OR = 1.69, 95% CI: 1.16–2.47, p = 0.006). Notably, home deliveries were associated with significantly reduced odds of low birthweight compared to deliveries at public health facilities (OR = 0.28, 95% CI: 0.14–0.59, p = 0.001). Deliveries in private facilities were linked to increased odds of low birthweight (OR = 1.61, 95% CI: 0.99–2.60), though this finding approached but did not reach statistical significance (p = 0.052). A particularly strong association was observed for multiple births, which were associated with an eight-fold increase in the odds of low birthweight compared to singleton births (OR = 8.05, 95% CI: 3.64–17.83, p < 0.001).

Model 2 was selected as the best-fitting model based on its lower Akaike Information Criterion (AIC) and Bayesian Information Criterion (BIC) values. Additionally, all variance inflation factors (VIF) values were below 5, indicating no evidence of problematic multicollinearity among the independent variables.

**Table 2:** Multilevel mixed effect logistic regression of low birthweight neonates of HIV-infected and non-infected adolescent and young adult mothers (n = 2,336)

| Variable | Model 1<br>OR | Model 2<br>OR | Model 3<br>OR |
| --- | --- | --- | --- |
| **Maternal HIV status** |  |  |  |
| HIV negative |  | ref (1) | ref (1) |
| HIV positive |  | 0.95 (0.50 - 1.79) | 0.93 (0.49 - 1.75) |
| **Maternal Age** |  |  |  |
| 15 to 19 |  | ref (1) | ref (1) |
| 20 to 24 |  | 0.78 (0.56 - 1.09) | 0.78 (0.56 - 1.09) |
| **Educational level** |  |  |  |
| None |  | ref (1) | ref (1) |
| Primary |  | 0.94 (0.48 - 1.84) | 0.95 (0.48 - 1.74) |
| Secondary |  | 0.63 (0.31 - 1.28) | 0.66 (0.32 - 1.36) |
| **Marital status** |  |  |  |
| Not married |  | ref (1) | ref (1) |
| Married |  | 0.86 (0.62 - 1.18) | 0.82 (0.59 - 1.15) |
| **Wealth status** |  |  |  |
| Poor |  | ref (1) | ref (1) |
| Middle |  | 1.69 (1.16 - 2.47)*** | 1.62 (1.10 - 2.38)** |
| Rich |  | 0.87 (0.50 - 1.51) | 0.82 (0.45 - 1.48) |
| **Employment status** |  |  |  |
| Not employed |  | ref (1) | ref (1) |
| Employed |  | 0.76 (0.55 - 1.05) | 0.76 (0.55 - 1.05) |
| **Media Exposure** |  |  |  |
| Not exposed |  | ref (1) | ref (1) |
| Exposed |  | 0.93 (0.67 - 1.28) | 0.85 (0.61 - 1.19) |
| **Residence** |  |  |  |
| Urban |  | ref (1) | ref (1) |
| Rural |  | 0.81 (0.54 - 1.21) | 0.78 (0.52 - 1.19) |
| **Improved water source** |  |  |  |
| Unimproved |  | ref (1) | ref (1) |
| Improved |  | 0.87 (0.62 - 1.21) | 0.79 (0.56 - 1.12) |
| **Place of delivery** |  |  |  |
| Public facility |  | ref (1) | ref (1) |
| Private facility |  | 1.61 (1.00 - 2.60) | 1.61 (0.99 - 2.61) |
| Home |  | 0.28 (0.14 - 0.59)*** | 0.30 (0.14 - 0.63)*** |
| **Multiple births** |  |  |  |
| Singleton |  | ref (1) | ref (1) |
| Multiple |  | 8.05 (1.00 - 2.60)*** | 9.11 (3.89 - 21.36)*** |
| **Antenatal care visits** |  |  |  |
| less than 8 |  |  | ref (1) |
| 8 or more |  |  | 0.92 (0.63 - 1.35) |
| **Province** |  |  |  |
| Central |  |  | ref (1) |
| Copperbelt |  |  | 1.07 (0.52 - 2.22) |
| Eastern |  |  | 1.25 (0.66 - 2.37) |
| Luapula |  |  | 1.34 (0.68 - 2.66) |
| Lusaka |  |  | 1.11 (0.53 - 2.31) |
| Muchinga |  |  | 0.85 (0.40 - 1.82) |
| Northern |  |  | 0.68 (0.31 - 1.50) |
| North western |  |  | 0.75 (0.36 - 1.54) |
| Southern |  |  | 1.01 (0.51 - 2.00) |
| Western |  |  | 0.79 (0.38 - 1.64) |
| AIC | 1983.778 | 1352.293 | 1364.837 |
| BIC | 1996.02 | 1438.635 | 1508.74 |
| VIF | < 5 | < 5 | > 5 |

\*\*\* = p < 0.001
\*\*\* = 0.05 < p < 0.01

## Discussion

This study investigated the effects of maternal HIV infection on low birthweight among children born to adolescent and young adult mothers in Zambia using data from the 2018 Zambia Demographic and health survey. Maternal HIV infection was not significant with low birth weight (LBW). This suggest that HIV infection alone may not be a primary determinant of LBW outcomes with the age group of adolescents and young women which is consistent with a study in Masvingo province [14]. The findings in this study are contrary to the ones conducted in Nigeria owari state in 2023, where it was revealed that HIV positive mothers had significant association with giving birth to babies with low birth weight [15]. A similarly in Tanzania HIV progression among mothers had increased risks of giving birth to babies with low birth weight [16].

A meta-analysis examining the relation of HIV infection to pregnancy outcomes concluded that HIV-infected women are at higher risk of LBW than are uninfected women [16,17]. The findings in this study revealed reduced odds between HIV status and LBW among children born to adolescent and young adult mothers can be due to the HIV interventions that Zambia has been implementing. Zambia has been implementing the UNAID 95 95 95 global goal which states that 95 percent of people living with HIV know their HIV status, 95 percent of people who know their HIV status are accessing treatment, and 95 percent of people on treatment have suppressed viral loads. The other interventions Zambia has implemented is PMTCT to prevent the transmission of HIV from the infected mother to the unborn baby, this coincides with a study conducted in Ethiopia which confirmed that improved counselling on LBW among pregnant women during ANC improved the outcome of the baby at birth [18].

Socioeconomic factors showed a significant association with low birth weight (LBW). Women in the middle wealth index were more likely to deliver LBW infants compared to poor women, while those in the rich wealth index had reduced odds of LBW albeit that it was not statistically significant. This finding contradicts with a study in Sylhet district, Bangladesh, which identified lower wealth status as a risk factor for LBW across all reproductive age groups[19]. Similarly, pooled demographic health surveys from 2005 to 2022 in low- and middle-income countries reported that infants born to poorer women had a higher likelihood of LBW compared to those born to wealthier mothers [20]. Wealth status is closely linked to food purchasing power and nutritional adequacy during pregnancy, with women of lower income more likely to face malnutrition and food insecurity[21]. These disparities, coupled with reduced access to quality healthcare, prenatal services, and increased psychosocial stress, contribute to adverse pregnancy outcomes and fetal growth restriction.

This study revealed that place of birth was significant predictor of LBW among adolescents and young women in Zambia. Infants born in private facilities had high likelihood of low birth weight compared to those delivered in public health facilities, these findings are consistent with a cohort study conduct at a public hospital of northwest of Ethiopia among HIV infected women[22]. In A study conducted in Ethiopia indicated that low birth weight was higher in public hospitals, 24.7% compared to private hospitals 15.2% [23]. The outcomes in the current study in relation to place of delivery could be linked to several factors. Private health facilities may be accessed by adolescents and young women who were seeking health servicers of higher socio-economic and demographic background with relatively poor health seeking behavior. Furthermore, there is a possibility that private health facilities receive complicated health issues from the public.

Multiple births showed the strongest association with LBW, with an eightfold higher risk compared to singletons. This was consistent with a multilevel multinomial regression analysis of low birth weight conducted in Sub-Saharan by Meklit and et al [24]. The findings in this study are also in line with the findings of a cross-sectional study that was conducted at Mulango Hospital in Uganda [25]. Maternal age, level of education, residence, marital status and antenatal care attendance did not show a significant association with low birth weight in our study. Contrary to a study conducted in Nigeria, Education level, antenatal care visits is a critical determinant of maternal and child health [2]. A study conducted in Ethiopia concluded a significant association between place of residence and LBW [26]. Findings in this study are supported by the study in Malawi were maternal age was not significantly associated with LBW in a multivariate analysis [27].

Eastern and Northern provinces reported the highest prevalence of LBW among HIV-infected adolescent mothers, while Muchinga and Luapula recorded the lowest rates. These findings suggest potential gaps in maternal and child health interventions, nutrition support, and adherence to HIV treatment protocols. These findings are supported by a study conducted in india although the study focused on general population of women [28]. In contrast, Muchinga and Luapula reported the lowest prevalence rates, which may reflect stronger program implementation, better health-seeking behaviors, or improved access to antenatal and HIV care services similar to a findings in southern Asia and Sub Saharan region [29]. These disparities highlight the need for tailored, province-specific strategies that address the unique challenges faced by adolescent mothers, particularly in high-burden areas, while reinforcing successful practices in regions with lower prevalence.

### Study Strength and study limitations

The strength of this study is that it focused on an important age group which is usually left out in the health care system. The sample size in this study might have affected the findings in this study. Another limitation is that the study did not consider the viral load suppression rates of adolescent’s girls and young women. Data that was used in this study was last collected in 2018.

### Policy recommendations

This study reveals that being HIV-positive does not increase the risk of giving birth to a low-birthweight baby among adolescents and young women in Zambia. Instead, factors including household income, place of delivery, multiple births seem to have a much stronger influence. Therefore, the following recommendations have been put up based on the findings in this study. Improve Maternal Nutrition and Support for Pregnant adolescents and young women. Young women need better access to healthy food and guidance on healthy eating. Community nutrition initiatives, supplements, and food support should target middle-income families, who had high risk of low birthweight infants.

Secondly, government should provide quality care accessible to both private and public health facilities through Regular quality checks, training for health workers, and stronger referral systems can help. Policies should also make it easier for young mothers to reach a facility with skilled birth attendants when they go into labour. Thirdly, more attention for multiple births should be given especially during ANC by health providers since being pregnant for more multiple babies increases the risk of LBW. Health providers should identify multiple pregnancies early during ANC visits and offer extra monitoring, nutritional advice, and specialized care to ensure safe and healthy deliveries.

The other policy recommendation is to focus on High-Burden Provinces such as Eastern, Northern, Copperbelt, and Northwestern which recorded higher LBW prevalence among HIV-positive mothers. More support should be directed to these areas, including improved maternal nutrition services, better HIV treatment follow-up, and more adolescent-friendly ANC services. Furthermore, continued use of community and media education Health information via radio, TV, or community gatherings improves outcomes for mothers. Public awareness initiatives can promote ANC attendance, nutrition, HIV medication adherence, and birth readiness. Special messaging for young mothers can reduce stigma and enhance health-seeking behaviour. Strengthen HIV programs although HIV status did not predict LBW in this study, PMTCT and the 95-95-95 approach must continue to improve. High testing, treatment, and virus suppression rates will help maintain favourable results.

## Conclusion

In conclusion, this study presents a strong argument for a transformative change in policymaking concerning maternal and child health in Zambia. Although the efficacy of HIV interventions is praiseworthy, an exclusive emphasis on HIV status may neglect other significant, amendable factors contributing to low birth weight (LBW). The results underscore the necessity for extensive, multi-sectoral interventions that are distinctly localized and attuned to provincial-level disparities. Future policies must prioritize improving maternal nutrition, ensuring fair access to high-quality prenatal care and delivery services, and offering specialized assistance to mothers expecting multiple births, especially among socioeconomically disadvantaged groups. Zambia can improve birth outcomes and protect the health and development of its youngest citizens by taking a more comprehensive approach that looks at the bigger picture of socioeconomic and maternal health.

## Data Availability

Data for this study 
were obtained from the Zambia Demographic 
and Health Survey (ZDHS), which is part of 
the Demographic and Health Surveys (DHS) 
Program. The data were originally accessed 
through the DHS Program website (https://
dhsprogram.com/).

## Acknowledgements

Thanks to the Zambia Statistical Agency (ZSA) and the DHS program for granting permission to utilize the 2018 ZDHS.

## References

1. Alam MJ, Islam MM, Maniruzzaman M, Ahmed NF, Tawabunnahar M, Rahman MJ, et al. Socioeconomic Inequality in the Prevalence of Low Birth Weight and Its Associated Determinants in Bangladesh. Plos one 2022, 17, e0276718.

2. Avwerhota O, Avwerhota M, Daniel E, Popoola T, Popoola I, Ogun A, et al. Determinants of Risk Factors Associated with Low Birth Weight in Nigeria. AJNHS 2024, 5, 77–87, doi:10.11648/j.ajnhs.20240503.15.

3. Natukunda P, Kadubira E, Nzabona A. Correlation between Maternal Factors and Birth Weight of Newborns in Uganda. 2024.

4. Mukosha M, Jacobs C, Kaonga P, Musonda P, Vwalika B, Lubeya MK, et al. Determinants and Outcomes of Low Birth Weight among Newborns at a Tertiary Hospital in Zambia: A Retrospective Cohort Study. Annals of African Medicine 2023, 22, 271–278.

5. 2021 Zambia Population Based HIV Impact Assessment, Final Report December 2023. https://www.zamstats.gov.zm/wp-content/uploads/2024/01/ZAMPHIA-2021-Final-Report.pdf.

6. Zambia Statistics Agency, Ministry of Health (MOH) Zambia, and ICF. 2019. Zambia Demographic and Health Survey 2018. Lusaka, Zambia, and Rockville, Maryland, USA: Zambia Statistics Agency, Ministry of Health, and ICF. https://dhsprogram.com/pubs/pdf/fr361/fr361.pdf

7. Bilal JA, Rayis DA, AlEed A, Al-Nafeesah A, Adam I. Maternal Undernutrition and Low Birth Weight in a Tertiary Hospital in Sudan: A Cross-Sectional Study. Frontiers in Pediatrics 2022, 10, 927518.

8. Minda B, Bekele G, Hailemeskel S, Lambebo A. Determinants of Low Birth Weight among Newborns Delivered in Public Hospitals of North Shewa Zone, Amhara Region, Ethiopia: A Case-Control Study (2023). Plos one 2024, 19, e0303364.

9. Echendu GE, Vincent CC, Ibebuike J, Asodike M, Naze N Chinedu EP, et al. Weights Of Infants Born to HIV Infected Mothers: A Prospective Cohort Study in Federal Medical Centre, Owerri, Imo State. European Journal of Pharmaceutical and Medical Research 2023, 10, 564–568.

10. Husaeni N, Cusmarih C. Meta-Analysis: The Effect of HIV/AIDS in Pregnant Women Against the Incidence of Low Birth Weight and Preterm Birth. In Proceedings of the Proceedings of the International Conference on Nursing and Health Sciences; 2023; Vol. 4, pp. 233–240.

11. Chola M, Michelo C. Proximate Determinants of Fertility in Zambia: Analysis of the 2007 Zambia Demographic and Health Survey. International Journal of Population Research 2016, 2016, 1–7, doi:10.1155/2016/5236351.

12. Mahumud RA, Sultana M, Sarker AR. Distribution and Determinants of Low Birth Weight in Developing Countries. Journal of preventive medicine and public health 2016, 50, 18.

13. Organization, W.H. WHO Recommendations on Antenatal Care for a Positive Pregnancy Experience: Summary: Highlights and Key Messages from the World Health Organization’s 2016 Global Recommendations for Routine Antenatal Care; World Health Organization, 2018;

14. Guvava V. Disparities in Birth Outcomes Between HIV-Positive Women Undergoing Antiretroviral Therapy and HIV-Negative Women in Masvingo Province, 2023-2024. 2025.

15. Echendu GE, Vincent CC, Ibebuike J, Asodike M, Naze N, Chinedu EP et al. Weights Of Infants Born to HIV Infected Mothers: A Prospective Cohort Study in Federal Medical Centre, Owerri, Imo State. European Journal of Pharmaceutical and Medical Research 2023, 10, 564–568.

16. Dreyfuss ML, Msamanga GI, Spiegelman D, Hunter DJ, Urassa EJ, Hertzmark E, et al. Determinants of Low Birth Weight among HIV-Infected Pregnant Women in Tanzania. The American journal of clinical nutrition 2001, 74, 814–826.

17. Brocklehurst P, French R. The Association between Maternal HIV Infection and Perinatal Outcome: A Systematic Review of the Literature and MetalJanalysis. BJOG 1998, 105, 836–848, doi:10.1111/j.1471-0528.1998.tb10227.x.

18. Mekonnen Y, Wolde E, Hagos T, Yared K, Bekele A, Mehari Z, et al. Improved Antenatal Care Services in Rural Ethiopia’s Public Health Centers through the Enhancing Nutrition and Antenatal Infection Treatment (ENAT) Intervention. BMC Health Serv Res 2025, 25, doi:10.1186/s12913-025-12789-4.

19. Bhattacharjee P, Islam MS, Roy A, Amin ZF, Rahman NB, Shahed MA et al. Incidence and Risk Factors of Low Birth Weight in a Rural District of Bangladesh: A Prospective Cohort Study. 2024.

20. Mare KU, Andarge GG, Sabo KG, Mohammed OA, Mohammed AA, Moloro AH et al. Regional and Sub-Regional Estimates of Low Birth Weight and Its Determinants in 44 Low- and Middle-Income Countries: Evidence from Demographic and Health Survey Data. BMC Pediatr 2025, 25, 342, doi:10.1186/s12887-025-05691-9.

21. Rohmah N, Masruroh M, Marasabesy NB, Pakaya N, Prasetyo J, Walid S, et al. Factors Related to Low Birth Weight in Indonesia. Malaysian Journal of Nutrition 2022, 28.

22. Kebede B, Andargie G, Gebeyehu A. Birth Outcome and Correlates of Low Birth Weight and Preterm Delivery among Infants Born to HIV-Infected Women in Public Hospitals of Northwest Ethiopia. Health 2013, 5, 25–34.

23. Chanie H, Dilie A. Prevalence of Low Birth Weight and Associated Factors among Women Delivered in Debre Markos Referral Hospital, East Gojam, Ethiopia, 2017. Prevalence 2018, 53, 1–10.

24. Bezie MM, Tesema GA, Seifu BL. Multilevel Multinomial Regression Analysis of Factors Associated with Birth Weight in Sub-Saharan Africa. Scientific Reports 2024, 14, 9210.

25. Louis B, Steven B, Margret N, Ronald N, Emmanuel L, Tadeo N et al. Prevalence and Factors Associated with Low Birth Weight among Teenage Mothers in New Mulago Hospital: A Cross Sectional Study. Journal of health science (El Monte) 2016, 4, 192.

26. Fentie EA, Yeshita HY, Bokie MM. Low Birth Weight and Associated Factors among HIV Positive and Negative Mothers Delivered in Northwest Amhara Region Referral Hospitals, Ethiopia, 2020 a Comparative Crossectional Study. PLoS One 2022, 17, e0263812.

27. Mfipa D, Hajison PL, Mpachika-Mfipa F. Predictors of Low Birthweight and Comparisons of Newborn Birthweights among Different Groups of Maternal Factors at Rev. John Chilembwe Hospital in Phalombe District, Malawi: A Retrospective Record Review. Plos one 2024, 19, e0291585.

28. Kundu RN, Ghosh A, Chhetri B, Saha, I.; Hossain, Md.G.; Bharati, P. Regional with Urban–Rural Variation in Low Birth Weight and Its Determinants of Indian Children: Findings from National Family Health Survey 5 Data. BMC Pregnancy Childbirth 2023, 23, 616, doi:10.1186/s12884-023-05934-6.

29. Okwaraji YB, Krasevec J, Bradley E, Conkle J, Stevens GA, Gatica-Domínguez G et al. National, Regional, and Global Estimates of Low Birthweight in 2020, with Trends from 2000: A Systematic Analysis. The Lancet 2024, 403, 1071–1080.

